# Genetic overlap and causal associations between smoking behaviours and psychiatric traits and disorders in adolescents and adults

**DOI:** 10.1101/2020.02.07.20021089

**Authors:** Wikus Barkhuizen, Frank Dudbridge, Angelica Ronald

## Abstract

**Background:** Epidemiological research shows that smoking is associated with psychiatric disorders and psychotic experiences, even after controlling for confounds such as cannabis use and sleep problems. We investigated degree of genetic overlap and tested for causal associations between smoking and psychiatric traits and disorders using genetic data. We tested whether genetic associations existed beyond genetic influences shared with confounding variables.

**Methods:** Genetic correlations were estimated with LD-score regression between smoking behaviours (N=262,990-632,802) and psychiatric disorders (schizophrenia, bipolar disorder and depression; N=41,653-173,005), psychotic experiences in adolescents (N=6,297-10,098) and adults (N=116,787-117,794) and adult schizotypy (N=3,967-4,057). Genomic Structural Equation Modelling was performed to explore the associations while accounting for genetic influences of confounders (cannabis and alcohol use, risk-taking and insomnia). Causal associations were tested using Generalized Summary-based Mendelian Randomization (GSMR).

**Results:** Significant genetic correlations were found between smoking and psychiatric disorders (r_g_ = .10 - .38) and adult PE (r_g_ = .33 - .40). After accounting for covarying genetic influences, genetic associations between most smoking phenotypes and schizophrenia and depression remained but not between smoking behaviours and bipolar disorder or most psychotic experiences. GSMR results supported a causal role of smoking initiation on psychiatric disorders and adolescent cognitive and negative psychotic experiences.

**Conclusions:** Pleiotropy between smoking behaviours and schizophrenia and depression exists beyond the common genetic basis of known confounders. Smoking also appears to be causally associated with psychiatric disorders and with cognitive PEs and negative symptoms during adolescence. Exploration of the biological links underlying smoking and psychiatric illness would be well-justified.

## Introduction

Despite declining smoking rates in high-income countries, smoking still affects a substantial proportion of the population, with 14-15% of adults in the US and UK and 28% across Europe smoking regularly (1-3). High co-occurrence between smoking behaviours and psychiatric disorders is well-established (4-6). Smoking rates among individuals diagnosed with schizophrenia or bipolar disorder are five times greater and with depression two-fold greater compared to healthy controls (7, 8). Smoking behaviours also co-occur with subthreshold psychiatric traits in the general population such as psychotic experiences (PE) (9-11). Regular smoking during adolescence has been associated with a range of psychotic experiences and negative symptom traits (PENS) (12).

In terms of underlying causes, a recent twin study found that regular smoking during adolescence shared genetic influences with paranoia and cognitive disorganization (r_A_=.37-.45), and familial influences with hallucinations (12). A previous study found no evidence that adolescent PENS was predicted by polygenic liability to initiate smoking (13), but used less well-powered genome-wide association study (GWAS) summary statistics than what are currently available. Schizophrenia and major depression share genome-wide genetic influences with smoking behaviours (14, 15), and polygenic liability to bipolar disorder has been associated with nicotine dependence (16).

There are several covariates that could, at a genetic level, account for potential genetic overlap between psychiatric traits or disorders and smoking behaviour. Epidemiological studies have investigated cannabis and alcohol use, stressful or traumatic life events, sociodemographic characteristics, novelty-seeking behaviour and sleep disturbances as confounders of the association between smoking behaviour and mental health problems (9-12, 17-21). Risk-taking behaviour have also been associated with smoking (22, 23) and psychiatric outcomes (24). Many of these covariates, including cannabis and alcohol use, risk-taking and insomnia, are partly under genetic influence and have been genetically associated with smoking or with psychiatric traits or disorders (25-30). This issue has not been addressed in studies to date.

Smoking behaviour may be a causal risk factor for psychiatric disorders based on evidence from longitudinal and dose-response associations and Mendelian randomization studies (17, 19, 21, 31, 32). Smoking behaviour may also be causally linked to PE. The association between PE and smoking is not fully explained by known confounding factors (9, 33-37, but see 33) and a dose-response relationship has been reported (9, 38) although not consistently (33, 36). Most longitudinal studies report an association between smoking behaviour and later reports of PE (10, 39-41) while evidence does not support the idea that adolescents start to smoke to alleviate pre-existing PE (39). However, these longitudinal associations do not necessarily reflect true causal associations because such study designs cannot completely rule out the possibility that PE were present prior to smoking initiation and vice-versa. Triangulation of these findings with evidence from methods such as Mendelian randomization is needed.

This study assessed the degree to which smoking behaviours are genetically correlated with PE across adolescence and adulthood and with schizophrenia, major depression and bipolar disorder. Second, we aimed to estimate the extent to which these genetic correlations remain after controlling for genetic influences on cannabis and alcohol use, risk taking behaviour and sleep disturbances (25-29). Third, we aimed to assess causal associations between smoking initiation and adult and adolescent PE and confirm previous reports of causal associations with psychiatric disorders.

## Methods

### Samples and measures

#### Smoking behaviours

Summary statistics for smoking initiation, cigarettes per day, age of initiation and current smoking (N = 262,990 - 632,802) were obtained from the GWAS and Sequencing Consortium of Alcohol and Nicotine use (GSCAN) (42) meta-GWAS on individuals of predominantly European ancestry. Smoking initiation was a binary phenotype with smokers defined as those who reported having ever smoked regularly. The average number of cigarettes per day was assessed in current and former smokers, with never-smokers excluded. Age of smoking initiation was defined as the age at which current or former smokers started smoking regularly. Current smoking was assessed among smokers and coded as either current smokers or former smokers.

#### Adolescent psychotic experiences and negative symptoms

Summary statistics were obtained from a mega-GWAS of four continuous scales of adolescent PENS among participants of European ancestry: Paranoia and hallucinations, cognitive disorganisation, parent-rated negative symptoms and anhedonia (N 6,297-10,098) (43). PENS items came from three community-based samples: The Twins Early Development Study (TEDS) (44) had a mean age of 16.32 years at the time of assessment, the Avon Longitudinal Study of Parents and Children (ALSPAC; mean age 16.76 years) (45, 46), and the Child and Adolescent Twin Study in Sweden (CATSS; mean age 18.31 years) (47).

Ethical approval for the original adolescent PENS GWAS (43) was obtained for ALSPAC from the ALSPAC Ethics and Law Committee and the Local Research Ethics Committees, for TEDS from the Institute of Psychiatry ethics committee (ref: 05/Q0706/228), and for CATSS from the Karolinska Institute Ethical Review Board. All research participants granted informed consent.

#### Schizotypy in adulthood

GWAS on four continuous schizotypy scales assessed during middle adulthood in the Northern Finland Birth Cohort 1996 (NFBC) (48) when participants were aged 31 years were obtained from the authors (N 3,967 – 4.057) (49). Perceptual aberrations were assessed using the Perceptual Aberration Scale (50), hypomania using the Hypomanic Personality Scale (51), social anhedonia with the Revised Social Anhedonia Scale and physical anhedonia using the Revised Physical Anhedonia Scale (52).

#### Psychotic experiences in adults

The presence of lifetime positive PE were assessed in the UK Biobank using four dichotomous items as part of a mental health questionnaire completed by 157,397 participants aged 40-69 years. Participants reported an average age of PE onset of 31.6 (s.d. = 17.6) years. Summary statistics for individuals of European ancestry were obtained from the Neale Lab (*http://www.nealelab.is/uk-biobank*) on experiences of auditory hallucinations, visual hallucinations, delusions of persecution and delusions of reference.

#### Psychiatric disorders

Summary statistics were obtained from the Psychiatric Genomics Consortium (https://www.med.unc.edu/pgc/results-and-downloads) meta-GWAS for schizophrenia (53) (N= 150,064), major depressive disorder (15) (N = 173,005 excluding 23andMe participants) and bipolar disorder (54) (N = 41,653). Diagnosis of schizophrenia was based on DSM-IV criteria for schizophrenia or schizoaffective disorder. Major depression diagnoses were based on clinical interviews, obtained from electronic healthcare records or based on self-report in some UK Biobank participants. Different clinical interview formats were used to diagnose Bipolar disorder, described in full elsewhere (54).

#### Covariates in genomic multiple regression

Publicly available summary statistics were obtained for lifetime cannabis use (N = 162,082) (30), alcohol consumption (N = 537,349 excluding 23andMe participants) (42), risk taking (Neale Lab; N = 348,549 UK Biobank participants) and insomnia (N= 113,006) (25).

Cannabis use was a binary phenotype assessed using self-report items on whether participants had ever used cannabis. Alcohol consumption came from participant reports on the average number of weekly drinks they drank. Risk taking was assessed with the item *“Would you describe yourself as someone who takes risks?”* (UK Biobank data-field 2040). Insomnia was from an item from the UK Biobank (data-field 1200) with participants who indicated that they usually have trouble falling asleep at night or wake up in the middle of the night classed as cases.

The Birkbeck Department of Psychological Sciences’ Ethics Committee approved this study.

### Analyses

GWAS summary statistics were filtered to remove single nucleotide polymorphisms (SNPs) with incomplete association statistics and to exclude strand ambiguous and non-biallelic SNPs. Variants were matched and allele orders harmonized to the 1000 Genomes (phase 3) reference panel for European ancestry. Variants were excluded based on INFO scores < 0.9 and minor allele frequency (MAF) < 0.01. INFO scores were not provided in the summary statistics for smoking behaviours and drinks per week. Imputed variants were filtered on INFO < 0.3 by the study authors (42). Details on summary statistics are provided in Supplementary Table 1.

#### LD score regression

Genetic correlations were estimated using linkage disequilibrium (LD) score regression (55, 56). SNP heritability estimates (SNP-h^2^) and genetic correlations (r_g_) were converted to a liability scale based on a population prevalence of 1% for schizophrenia, 15% for major depression and 2% for bipolar disorder (57-59). The genetic covariance intercept was left unconstrained in analyses with overlapping samples (GWASs for smoking behaviours, depression and adult positive PE included UK Biobank participants; GWASs for smoking behaviours and adolescent PENS included ALSPAC participants). Correction for multiple testing of genetic correlations (60 tests) was performed using Benjamini-Hochberg correction at a false discovery rate (FDR) of 0.05.

#### Genomic structural equation modelling

To investigate genetic overlap between psychiatric phenotypes and smoking behaviours after accounting for the genetic influences on confounds, Genomic Structural Equation Modelling (Genomic SEM) was used (60). Genomic SEM uses summary statistics to model the shared genetic architecture of genetically correlated traits using the genetic covariance structures.

LD structure for all phenotypes in the models was estimated in LD score regression software using the same parameters described above. Genomic covariance structures were computed, and genomic multiple regression models specified in the Genomic SEM package (60) for R version 3.5.2 (61) for PE and psychiatric disorders that had at least nominally significant genetic correlations (at *p* < .05) with smoking phenotypes. Models allowed for covariation between the genetic components of each smoking phenotype plus the covariates as predictors regressed onto a psychiatric outcome. Standardized estimates were reported thereby allowing the association between a given predictor and the outcome to be interpreted as genetic correlations conditional on all other predictors. Model fit was evaluated based on the Standardized Root Mean Square Residual (SRMR) statistic with SRMR < .05 considered a good fit.

#### Mendelian randomization

Mendelian randomization (MR) (62) was performed to test for bi-directional causal associations between smoking initiation and psychiatric phenotypes using summary statistics.

Generalised Summary-data-based Mendelian Randomisation (GSMR; Zhu et al., 2018) was used as the main MR method due to its advantages of accounting for sampling variation in the exposure and outcome GWAS and for residual LD structure between variants used as instrumental variables (IVs). MR-Egger regression (64), Weighted Median (65) and Weighted Mode MR (66) were conducted as sensitivity analyses as these methods make different assumptions to GSMR by allowing for a proportion of invalid IVs in the analyses.

The presence of directional pleiotropy was assessed using the MR-Egger intercept test. An intercept significantly different from zero indicates that MR-Egger causal estimates may be more robust compared to GSMR estimates.

Summary statistics for major depression excluded 23andMe participants and did not have enough variants at genome-wide significance. Genome-wide significant variants to use as IVs were instead obtained from the publication (15). Only seven variants reached genome-wide significance in the publicly available summary statistics for bipolar disorder (54). IVs for bipolar disorder were instead obtained from a recent GWAS for which full summary statistics were not available (67). For all other exposure phenotypes, IVs were identified using the clumping algorithm in PLINK (68) based on an r^2^ threshold = .05 within a 500kb window. Recent PE GWAS have not yet been replicated using equivalent measures in independent samples or are based on relatively small sample sizes. Thus, IVs for PE phenotypes were selected at *p* < 5 × 10^−5^.

Analyses were performed in GSMR (Zhu et al., 2018) and MR Base (69) packages for R version 3.5.2 (R Core Team, 2018). GCTA (70) was used to calculate the LD structure between lead variants based on the 1000 Genomes (phase 3) reference panel for European ancestry. IVs excluded from GSMR analyses due to being Heidi-outliers and in residual LD at an r^2^ threshold of 0.1 were also removed prior to conducting MR sensitivity analyses. MR was conducted using summary statistics for smoking initiation but not for smoking phenotypes assessed in smokers only. This is because genome-wide significant associations among smokers may not explain smoking liability in samples that include non-smokers. Significance thresholds were set at *p* < 0.05.

## Results

### Genetic overlap between smoking behaviours and psychiatric traits/disorders

Genetic correlations between smoking behaviours and psychotic experiences and psychiatric disorders are summarised in Figure 1.

**Figure 1.**
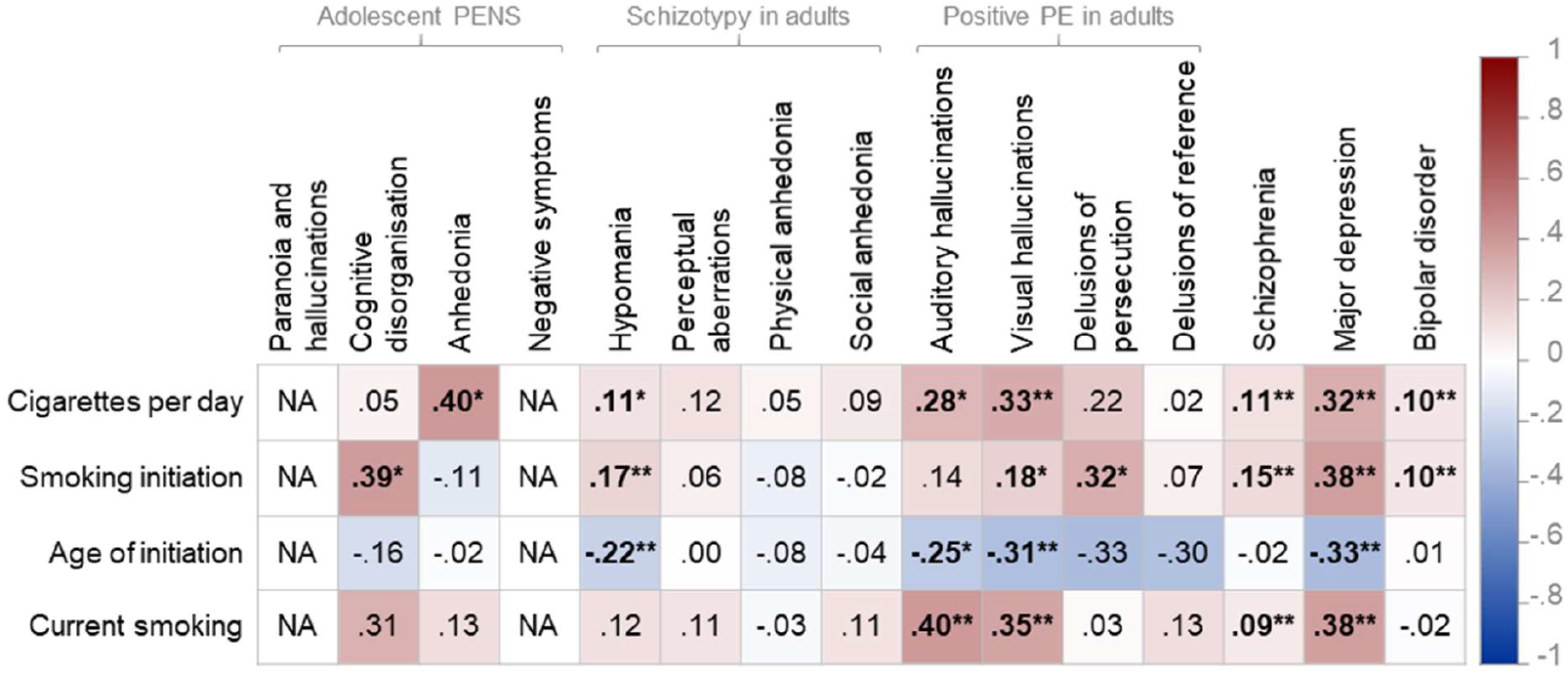
Heat map showing genetic correlations between smoking behaviours, psychotic experiences and psychiatric disorders. *Note:* PENS = Psychotic experiences (PE) and negative symptom traits; NA = indicates that genetic correlations could not be computed due to low SNP-heritability or sample size; * indicates statistically significant genetic correlations at p<.05; ** indicates significance at FDR < .05 using Benjamini-Hochberg correction for 60 tests; Genetic correlations reported using unconstrained LD score regression intercept between phenotypes with sample overlap (for example, smoking behaviour and major depression GWASs contained participants from the UK Biobank, and smoking behaviours and adolescent PENS contained participants from ALSPAC). Note that age of smoking initiation was coded in the direction of lower scores reflecting younger ages of initiation. Note that current smoking cases are current smokers (compared to ex-smokers).

For positive PE in adults, significant genetic correlations after correction for multiple testing were observed between experiences of visual hallucinations with cigarettes per day (r_g_ = .33, *p* = 4.00 × 10^−4^), age of smoking initiation (r_g_ = -.31, *p* = 0.002), current smoking (r_g_ = .35, *p* = 0.001), and nominally at *p* <.05 with smoking initiation (r_g_ = .18, *p* = 0.026).

Experiences of auditory hallucinations was significantly genetically correlated with current smoking (r_g_ = .40, *p* = 0.009), and nominally significant with cigarettes per day (r_g_ = .28, *p* = 0.015) and age of smoking initiation (r_g_ = -.25, p = 0.037). Delusions of persecution was genetically correlated at nominal significance with smoking initiation (r_g_ = .32, *p* = 0.016).

Hypomania was the only schizotypy measure to genetically correlate with smoking behaviours, specifically with smoking initiation (r_g_ = .17, *p* = 8.00 × 10^−4^), age of smoking initiation (r_g_ = -.22, *p* = 0.002) and at nominal significance with cigarettes per day (r_g_ = .11, *p* = 0.049).

For adolescent PENS, nominally significant genetic correlations were found between cognitive disorganisation and smoking initiation (r_g_ = .39, *p* = 0.017) and between anhedonia and cigarettes per day (r_g_ = .40, *p* = 0.046). We could not estimate genetic correlations for paranoia and hallucinations and negative symptoms due to the low SNP-h^2^. To indicate presence and direction of genetic overlap, we instead report genetic covariances. After correction for multiple testing, significant genetic covariation in the expected directions were found between paranoia and hallucinations with smoking initiation (ρ_g_ = 0.02, *p* = 0.010) and age of smoking initiation (ρ_g_ = -0.03, *p* = 1.16 × 10^−4^), and between negative symptoms and smoking initiation (ρ_g_ = 0.03, *p* = 0.001). Genetic covariation was also suggested (at *p* < .05) between negative symptoms with cigarettes per day (ρ_g_ = 0.02, *p* = 0.046) and with current smoking (ρ_g_ = 0.02, *p* < 0.050).

As expected, we found significant genetic correlations between smoking behaviours and psychiatric disorders (42). Schizophrenia was genetically correlated with cigarettes per day (r_g_ = .11, *p* = 2.00 × 10^−4^), smoking initiation (r_g_ = .15, *p* = 9.51 × 10^−11^) and current smoking (r_g_ = .09, *p* = 0.008). Major depression shared genetic influences with cigarettes per day (r_g_ = .32, *p* = 6.15 × 10^−12^), smoking initiation (r_g_ = .38, *p* = 1.57 × 10^−29^), age of smoking initiation (r_g_ = -.33, *p* = 1.16 × 10^−14^) and current smoking (r_g_ = .38, *p* = 1.31 × 10^−14^). Significant genetic correlations were found between bipolar disorder and cigarettes per day (r_g_ = .10, *p* = 0.006) and smoking initiation (r_g_ = .10, *p* = 7.00 × 10^−4^).

### Genomic multiple regression

To investigate the degree to which genetic correlations between smoking behaviours and psychiatric traits and disorders exist beyond genetic influences associated with covariates (lifetime cannabis use, alcohol consumption, insomnia and risk-taking behaviour), genomic multiple regression models were run in Genomic SEM for phenotype pairs that shared at least nominally significant (*p* < .05) genetic overlap (presented above and in Figure 1). Genetic correlations with the covariates are shown in Supplementary Figure 1.

All models provided excellent fit to the data (maximum SRMR = 8.30 × 10^−9^). Figure 2 shows the path diagram between schizophrenia and cigarettes per day as an example of the genomic multiple regression models (see Supplementary Figures 2-5 for path diagrams for all models). Table 1 summarizes the conditional genetic correlations (*b*_g_) between smoking behaviours and psychiatric disorders/PE obtained from these models.

**Table 1.** Conditional genetic correlations after accounting for genetic influences on covariates in genomic structural equation models

| Psychotic disorders | Smoking behaviours | LD score regression | Standardized conditional genetic associations |  |  |
| --- | --- | --- | --- | --- | --- |
|  |  | r<sub>g</sub> | b<sub>g</sub> | SE | p |
| Schizophrenia | Cigarettes per day | 0.11 | 0.12 | 0.04 | 0.003 |
| Schizophrenia | Smoking initiation | 0.15 | 0.00 | 0.04 | 0.970 |
| Schizophrenia | Current smoking | 0.09 | 0.12 | 0.05 | 0.009 |
| Major depression | Cigarettes per day | 0.32 | 0.17 | 0.06 | 0.003 |
| Major depression | Smoking initiation | 0.38 | 0.16 | 0.07 | 0.017 |
| Major depression | Age of smoking initiation | -0.33 | -0.11 | 0.06 | 0.075 |
| Major depression | Current smoking | 0.39 | 0.30 | 0.07 | 1.93 x 10<sup>-5</sup> |
| Bipolar disorder | Cigarettes per day | 0.10 | 0.05 | 0.05 | 0.263 |
| Bipolar disorder | Smoking initiation | 0.10 | -0.11 | 0.05 | 0.029 |
| Adult PE |  |  |  |  |  |
| Auditory hallucinations | Cigarettes per day | 0.28 | 0.05 | 0.12 | 0.662 |
| Auditory hallucinations | Age of smoking initiation | -0.25 | 0.00 | 0.12 | 0.999 |
| Auditory hallucinations | Current smoking | 0.40 | 0.21 | 0.14 | 0.125 |
| Visual hallucinations | Cigarettes per day | 0.33 | 0.18 | 0.13 | 0.165 |
| Visual hallucinations | Smoking initiation | 0.18 | 0.14 | 0.12 | 0.266 |
| Visual hallucinations | Age of smoking initiation | -0.31 | -0.15 | 0.13 | 0.248 |
| Visual hallucinations | Current smoking | 0.35 | 0.26 | 0.13 | 0.044 |
| Delusions of persecution | Smoking initiation | 0.32 | 0.20 | 0.18 | 0.258 |
| Schizotypy |  |  |  |  |  |
| Hypomania | Cigarettes per day | 0.11 | -0.01 | 0.17 | 0.968 |
| Hypomania | Smoking initiation | 0.17 | 0.00 | 0.16 | 0.980 |
| Hypomania | Age of smoking initiation | -0.22 | -0.04 | 0.16 | 0.820 |
| Adolescent PENS |  |  |  |  |  |
| Anhedonia | Cigarettes per day | 0.40 | 0.44 | 0.22 | 0.044 |
| Cognitive disorganization | Smoking initiation | 0.39 | 0.09 | 0.22 | 0.676 |

Note: $r_g$ = Bivariate genetic correlation estimates calculated in LD score regression; $b_g$ = Conditional genetic association estimates obtained from Genomic Structural Equation Modelling that, in standardized form, can be interpreted as conditional genetic correlations accounting for genetic covariation between predictors and regression paths of covariates onto psychiatric outcomes; PE = Psychotic experiences; PENS = Psychotic experiences and negative symptom traits.

**Figure 2.**
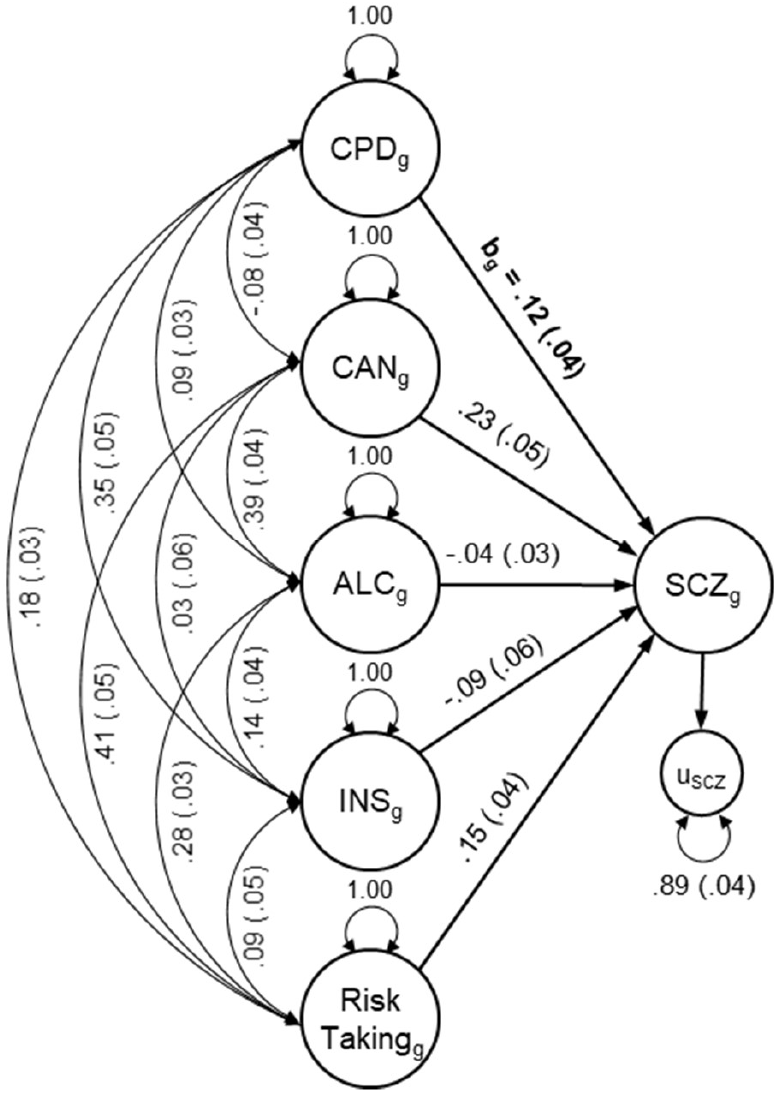
Path diagram illustrating genetic multiple regression models. *Note:* CPDg = Genetic component of cigarettes per day; CANg = Genetic component of cannabis use; ALCg = Genetic component of alcohol consumption; INSg = Genetic component of insomnia; SCZg = Genetic component of schizophrenia; u = residual genetic variance; bg = Conditional genetic correlation between the genetic components of cigarettes per day and schizophrenia as summarized in Table 1; Double-headed arrows represents genetic covariance; Single-headed arrows represents regression paths; Path diagrams for models between all smoking behaviours and psychiatric phenotypes are provided in Supplementary Figures 2-5.

The genetic component of cigarettes per day, plus the genetic influences on the four covariates as predictors, accounted for 11-41% of genetic variation in psychiatric traits and disorders (calculated as one minus the residual variance). Models using the genetic component of smoking initiation accounted for 10-52%, using age of smoking initiation 24-41%, and using current smoking status 11-41% of genetic variation in psychiatric traits and disorders.

Table 1 shows that after accounting for the genetic influences of the other covariates, conditional genetic associations (b_g_) were significant between cigarettes per day and schizophrenia (unattenuated compared to r_g_ estimates from LD score regression), depression (with b_g_ accounting for 53% of r_g_; calculated as b_g_/r_g_ × 100) and adolescent anhedonia (unattenuated) but not with bipolar disorder, auditory hallucinations, visual hallucinations and hypomania.

Significant conditional genetic associations were found between smoking initiation and depression (accounting for 42% of the r_g_ estimate from LD score regression) and a negative conditional genetic association with bipolar disorder (whereas r_g_ from LD score regression was positive). No genetic association was found between smoking initiation and schizophrenia, visual hallucinations, delusions of persecution, hypomania and adolescent cognitive disorganization after accounting for the genetic influences of covariates.

No significant conditional genetic associations were found between age of smoking initiation with depression, auditory hallucinations, visual hallucinations and hypomania. Negative conditional genetic association estimates indicate an association with a younger age of smoking initiation.

Significant genetic overlap not accounted for by other predictors in the model was found between current smoking and schizophrenia (with b_g_ being slightly higher than the r_g_ estimate from LD score regression), depression (accounting for 79% of r_g_ from LD score regression) and visual hallucinations (74% of r_g_ from LD score regression) and not with auditory hallucinations.

### Mendelian randomization

Table 2 presents MR results (see also Supplementary Figures 6-13 and Supplementary Table 2). GSMR analyses suggested a causal association between smoking initiation on schizophrenia liability with this finding replicated in Weighted Median but not in MR-Egger or Weighted Mode sensitivity analyses. A significant but smaller causal effect of schizophrenia liability on smoking propensity was found.

**Table 2.** Mendelian Randomization results.

| Exposure | Outcome | Heidi<br>SNPs | LD<br>SNPs | n<br>SNP | GSMR results |  |  | MR-Egger |  |  | Weighted Median |  |  | Weighted Mode |  |  |
| --- | --- | --- | --- | --- | --- | --- | --- | --- | --- | --- | --- | --- | --- | --- | --- | --- |
|  |  |  |  |  | Beta | SE | p | Beta | SE | p | Beta | SE | P | Beta | SE | p |
| Smoking initiation | → Schizophrenia | 9 | 3 | 102 | 0.653 | 0.113 | **6.89 x 10<sup>-9</sup>** | 0.022 | 0.900 | 0.981 | 0.496 | 0.192 | **0.010** | -0.372 | 0.812 | 0.647 |
| Schizophrenia | → Smoking initiation | 12 | 11 | 81 | 0.011 | 0.003 | **4.12 x 10<sup>-4</sup>** | 0.016 | 0.019 | 0.398 | 0.010 | 0.005 | **0.031** | -0.006 | 0.012 | 0.630 |
| Smoking initiation | → Major depression | 6 | 2 | 103 | 0.854 | 0.083 | **1.51 x 10<sup>-24</sup>** | 0.834 | 0.516 | 0.109 | 0.809 | 0.129 | **3.84 x 10<sup>-10</sup>** | 0.852 | 0.297 | **0.005** |
| Major depression | → Smoking initiation | 4 | 0 | 28 | 0.000 | 0.000 | 0.638 | 0.073 | 0.015 | **5.34 x 10<sup>-5</sup>** | 0.000 | 0.001 | 0.611 | -0.001 | 0.002 | 0.761 |
| Smoking initiation | → Bipolar disorder | 4 | 3 | 106 | 0.812 | 0.151 | **7.82 x 10<sup>-8</sup>** | 0.795 | 0.955 | 0.407 | 0.800 | 0.232 | **0.001** | 0.893 | 0.542 | 0.102 |
| Bipolar disorder | → Smoking initiation | 0 | 0 | 18 | 0.006 | 0.007 | 0.356 | -0.098 | 0.058 | 0.112 | -0.003 | 0.010 | 0.755 | -0.009 | 0.016 | 0.562 |
| Smoking initiation | → Auditory hallucinations | 0 | 3 | 116 | 0.003 | 0.005 | 0.496 | 0.013 | 0.023 | 0.573 | 0.002 | 0.007 | 0.761 | 0.013 | 0.020 | 0.527 |
| Auditory hallucinations | → Smoking initiation | 0 | 0 | 89 | 0.041 | 0.085 | 0.631 | 0.297 | 0.186 | 0.113 | 0.119 | 0.117 | 0.310 | 0.197 | 0.334 | 0.557 |
| Smoking initiation | → Visual hallucinations | 1 | 3 | 115 | 0.010 | 0.007 | 0.169 | 0.030 | 0.033 | 0.358 | 0.014 | 0.010 | 0.172 | -0.007 | 0.024 | 0.781 |
| Visual hallucinations | → Smoking initiation | 0 | 0 | 81 | 0.043 | 0.066 | 0.513 | 0.128 | 0.154 | 0.408 | -0.005 | 0.089 | 0.958 | -0.205 | 0.225 | 0.365 |
| Smoking initiation | → Delusions of reference | 0 | 3 | 116 | 0.003 | 0.003 | 0.325 | 0.026 | 0.015 | 0.077 | 0.004 | 0.005 | 0.454 | 0.008 | 0.013 | 0.568 |
| Delusions of reference | → Smoking initiation | 0 | 0 | 105 | 0.005 | 0.125 | 0.970 | -0.036 | 0.259 | 0.890 | -0.062 | 0.173 | 0.722 | -0.296 | 0.458 | 0.520 |
| Smoking initiation | → Delusions of persecution | 0 | 3 | 116 | 0.005 | 0.003 | 0.171 | 0.008 | 0.015 | 0.624 | 0.002 | 0.005 | 0.688 | 0.000 | 0.012 | 0.995 |
| Delusions of persecution | → Smoking initiation | 0 | 0 | 92 | 0.035 | 0.126 | 0.778 | 0.137 | 0.309 | 0.657 | 0.027 | 0.182 | 0.882 | 0.212 | 0.462 | 0.647 |
| Smoking initiation | → Hypomania | 0 | 3 | 98 | 0.235 | 0.234 | 0.316 | 0.069 | 1.120 | 0.951 | 0.111 | 0.358 | 0.757 | -0.040 | 0.841 | 0.962 |
| Hypomania | → Smoking initiation | 0 | 0 | 68 | -0.001 | 0.002 | 0.565 | -0.004 | 0.005 | 0.494 | -0.002 | 0.003 | 0.461 | -0.003 | 0.006 | 0.577 |
| Smoking initiation | → Perceptual aberrations | 0 | 3 | 98 | 0.036 | 0.232 | 0.878 | 0.591 | 1.168 | 0.614 | 0.154 | 0.339 | 0.649 | 0.591 | 1.168 | 0.614 |
| Perceptual aberrations | → Smoking initiation | 0 | 0 | 54 | 0.000 | 0.002 | 0.867 | -0.001 | 0.005 | 0.792 | -0.001 | 0.003 | 0.732 | 0.000 | 0.006 | 0.953 |
| Smoking initiation | → Physical anhedonia | 0 | 3 | 98 | -0.139 | 0.234 | 0.553 | -0.464 | 1.201 | 0.700 | -0.170 | 0.360 | 0.636 | -0.180 | 0.788 | 0.820 |
| Physical anhedonia | → Smoking initiation | 1 | 0 | 58 | 0.000 | 0.002 | 0.850 | 0.003 | 0.006 | 0.626 | 0.002 | 0.003 | 0.524 | 0.010 | 0.008 | 0.209 |
| Smoking initiation | → Social anhedonia | 0 | 3 | 98 | -0.414 | 0.233 | 0.076 | -0.579 | 1.118 | 0.605 | -0.469 | 0.347 | 0.177 | -0.326 | 0.861 | 0.706 |
| Social anhedonia | → Smoking initiation | 1 | 0 | 57 | -0.001 | 0.002 | 0.754 | 0.005 | 0.005 | 0.294 | 0.002 | 0.003 | 0.578 | 0.004 | 0.006 | 0.530 |
| Smoking initiation | → Paranoia/hallucinations | 0 | 1 | 57 | 0.282 | 0.213 | 0.185 | 0.437 | 0.997 | 0.663 | 0.073 | 0.296 | 0.804 | -0.056 | 0.564 | 0.922 |
| Paranoia/hallucinations | → Smoking initiation | 0 | 0 | 24 | -0.002 | 0.006 | 0.720 | -0.015 | 0.015 | 0.330 | -0.006 | 0.008 | 0.460 | -0.015 | 0.014 | 0.322 |
| Smoking initiation | → Cognitive disorganisation | 0 | 1 | 57 | 1.030 | 0.261 | **8.02 x 10<sup>-5</sup>** | 2.321 | 1.191 | 0.056 | 1.267 | 0.365 | **5.25 x 10<sup>-4</sup>** | 1.352 | 0.691 | 0.055 |
| Cognitive disorganisation | → Smoking initiation | 0 | 0 | 28 | 0.004 | 0.004 | 0.307 | -0.022 | 0.011 | 0.059 | 0.002 | 0.006 | 0.723 | -0.001 | 0.010 | 0.943 |
| Smoking initiation | → Anhedonia | 0 | 1 | 57 | 0.058 | 0.244 | 0.812 | -0.841 | 1.208 | 0.490 | -0.075 | 0.362 | 0.835 | -0.218 | 0.604 | 0.720 |
| Anhedonia | → Smoking initiation | 0 | 0 | 30 | -0.002 | 0.004 | 0.599 | 0.000 | 0.009 | 0.965 | 0.000 | 0.006 | 0.944 | 0.002 | 0.011 | 0.868 |
| Smoking initiation | → Negative symptoms | 0 | 1 | 57 | 0.506 | 0.204 | **0.013** | 1.221 | 1.089 | 0.267 | 0.450 | 0.302 | 0.136 | 0.529 | 0.533 | 0.326 |
| Negative symptoms | → Smoking initiation | 0 | 0 | 25 | -0.003 | 0.006 | 0.609 | -0.012 | 0.015 | 0.436 | -0.004 | 0.008 | 0.664 | 0.004 | 0.015 | 0.780 |

Note: GSMR = Generalized Summary-based Mendelian Randomization; LD SNPs = SNPs with residual LD at $r^2 > 0.1$ removed from analysis; n SNPs = number of variants remaining in analyses after those identified as Heidi-outliers or with residual LD were removed. SNPs identified as having residual LD and as Heidi outliers were also excluded from MR Egger, Weighted Median and Weighted Mode analyses. Bold text indicates significant p-values at $p < 0.05$ .

Evidence of a causal association between smoking initiation and liability to major depression was found and a similar effect size was observed in MR-Egger (but with larger standard error), and replicated in Weighted Median and Weighted Mode MR. MR-Egger indicated that depression liability had a small but significant causal effect on smoking initiation. While this effect was not found in the other MR methods, the MR-Egger intercept was significantly different from zero (*p* = 5.53 × 10^−5^) suggesting the presence of directional pleiotropy, in which case the MR-Egger estimate is likely to be less biased than those of the other methods.

Evidence of a causal association between smoking initiation on liability for bipolar disorder was reported in GSMR and Weighted Median analyses with a similar effect size observed in MR-Egger, but not replicated in the Weighted Mode analysis. No evidence of reverse causation was found.

Some evidence of a causal effect of smoking initiation was observed for cognitive disorganisation in GSMR and Weighted Median analyses but was not replicated in MR-Egger and Weighted Mode MR, and on negative symptoms in the main MR analyses but not replicated in MR sensitivity analyses. No evidence of reverse causation was observed.

## Discussion

Smoking is the leading cause of preventable deaths and smoking rates are modifiable: they can be reduced through population-based interventions. We found evidence of overlapping genome-wide genetic influences between smoking behaviours and psychiatric disorders as well as between smoking behaviours and specific types of PE and negative symptom traits reported by adolescents and adults in the community. We further investigated the nature of pleiotropy between smoking behaviours and psychiatric traits and disorders. For schizophrenia and depression, overlap in common genetic influences associated with smoking frequency and current smoking status was not fully explained by genetic influences on known covariates, namely cannabis use, alcohol use, risk taking and insomnia. Genetic associations between smoking behaviours and most positive PE were explained by genetic influences shared with the covariates. We found evidence of causal effects of smoking initiation on schizophrenia, depression and bipolar disorder, as well as on adolescent cognitive and (to some degree) negative symptoms.

Our findings support the hypothesis that schizophrenia and depression share genetic pathways with smoking frequency and persistent smoking. Possible shared biological pathways may involve nicotine, the principal pharmacologically active component of smoking that acts as an agonist on the nicotinic acetylcholine receptor (nAChR). Variants within the CHRNA5-A3-B4 gene cluster which encodes for subunits of nAChR are among the most robust associations with nicotine dependence (42, 71-73) and have also been implicated in schizophrenia (53). Presynaptic activation of nAChR stimulates the release of several neurotransmitters including dopamine, serotonin and glutamate (74-77). Dysregulation of dopaminergic and glutamatergic pathways could both explain why some people may be more susceptible to the positive reinforcing effects of smoking (78) and have an increased vulnerability to develop schizophrenia (79).

We found support for a causal role of smoking initiation on liabilities to develop schizophrenia, depression and bipolar disorder, similar to the findings from two recent studies (31, 32) whilst using different Mendelian randomization methods. Our study took steps to remove likely pleiotropic variants from genetic instruments which aims to reduce confounding from biological pleiotropy. Our results of a possible causal effect of smoking on schizophrenia and depression (and a possible weaker effect in the other direction) concurs with meta-analytic findings of longitudinal studies (19, 21).

The action of nicotine on nAChR could also explain possible mechanisms by which smoking could be causally associated with psychiatric disorders. Chronic exposure to nicotine may result in long-lasting alterations of dopaminergic and cholinergic pathways, leading to an increase in risk of psychiatric disorders (80-82). Beyond nicotine, other toxic compounds released during combustion of tobacco cause neuro-inflammation and oxidative stress (Goncalves et al., 2011), factors that are associated with psychiatric disorders (Berk et al., 2011; Howes & McCutcheon, 2017; Miller, Maletic, & Raison, 2009).

To our knowledge, this is the first study to report that genome-wide genetic influences on bipolar disorder significantly overlap with those on smoking frequency and initiation. This finding supports recent evidence of an association between polygenic scores for bipolar disorder and nicotine dependence (16). Genetic overlap between bipolar disorder and smoking quantity was accounted for by genetic influences on the covariates. Interestingly, we found that smoking initiation was associated with lower genetic liability to be diagnosed with bipolar disorder, while the bivariate genetic correlation between these two phenotypes was positive. The negative conditional genetic association was likely due to smoking initiation being genetically correlated with all covariates (r_g_ = .25-.51).

Until recently, research into the genetic etiology underlying the association between PE in the community and smoking behaviours was lacking. Here, we found novel evidence that smoking behaviours share genetic influences with some types of positive PE during adulthood (notably with visual and auditory hallucinations) and with hypomania. Interestingly, for most associations, the genetic overlap between PE and smoking was shared with the covariates. We found suggestive evidence (p < .05) that adolescent anhedonia was genetically associated with cigarettes per day even after accounting for the genetic influences on confounds.

Smoking may increase a propensity to experience cognitive and negative PE during adolescence, although the association with negative symptom traits were not confirmed by Mendelian randomization sensitivity analyses. A recent preclinical study found that during adolescence, exposure to nicotine could lead to persistent alteration of neurotransmitter pathways (81) likely relevant to PE. Another possible causal mechanism is the effect of nitrogen oxides: Nitrogen oxide levels in air pollution was recently linked to an increased risk of PE in adolescents (83) and it is a toxin produced during cigarette smoking.

We replicated previous findings of genetic overlap between smoking behaviours and schizophrenia and major depression using more recent and better powered GWAS results (14, 15). We found a slightly higher genetic correlation of .38 between major depression and smoking initiation than the value of .29 reported by Wray et al. (15) and genetic correlations of similar magnitude with the other smoking phenotypes.

Smoking initiation and positive PE in adulthood did not appear to be causally related. This is somewhat surprising given the known phenotypic association between PE and psychiatric disorders (84-89) and could be explored further. Evidence suggests PE in adults have somewhat different underlying causal influences to PE during adolescence (90) and this may explain that lack of causal associations of PE in later life with smoking behaviours.

This study had limitations that should be considered. Our findings on schizotypy and adolescent PENS were limited by the relatively small GWAS sample sizes. As such, in some genomic multiple regression models the standard errors of the residual variances were large. We were unable to include some additional potential confounds such as exposure to trauma and sociodemographic characteristics. Large biobanks include biases from ascertainment and attrition. However, we note that attrition based on phenotypic or genetic risk for mental health conditions would lead us, if anything, to underestimate the genetic overlap between psychiatric disorders and traits with smoking behaviours in our analyses.

## Data Availability

This manuscript used genome-wide association study (GWAS) summary statistics, most of which is available online from:
https://genome.psych.umn.edu/index.php/GSCAN
https://www.med.unc.edu/pgc/data-index/
Summary statistics for adolescent psychotic experiences and for schizotypy may be requested from the authors of the original GWAS and requires access permission from ALSPAC and NFBB.

## Acknowledgements

This work was supported by the UK Medical Research Council (G1100559 to AR) and a Wellcome Trust ISSF grant (204770/Z/16/Z) and the Camara-Rijvers David Studentship to WB.

The authors gratefully acknowledge the ongoing contribution of the participants and their families in the Twins Early Development Study (TEDS), the Avon Longitudinal Study of Children and Parents (ALSPAC), the Child and Adolescent Twin Study in Sweden (CATSS) and participants in the North Finland Birth Cohort (NFBC) and the UK Biobank. We thank the funding bodies and research teams which includes interviewers, computer and laboratory technicians, clerical workers, research scientists, volunteers, managers, receptionists and nurses. The data from TEDS was supported by a program grant to Robert Plomin from the UK Medical Research Council (MR/M021475/1) and UK Medical Research Council grant G1100559 to AR. The UK Medical Research Council and Wellcome (Grant ref: 102215/2/13/2) and the University of Bristol provide core support for ALSPAC. This publication is the work of the authors and WB and AR will serve as guarantors for its contents. ALSPAC GWAS data was generated by Sample Logistics and Genotyping Facilities at Wellcome Sanger Institute and LabCorp (Laboratory Corporation of America) using support from 23andMe. We thank the Neale Lab and the Psychiatric Genomics Consortium (PGC) for providing genetic summary results and Dr. W. Hennah and A. Ortega-Alonso for preparing and sharing the schizotypy summary statistics. The authors are very grateful to Andrew Grotzinger for his advice on the implementation and interpretation of genomic multiple regression models performed within the Genomic Structural Equation Modelling software.

## Notes

### Competing Interest Statement

The authors have declared no competing interest.

